# Polygenic modifiers of expressivity in telomere biology disorders

**DOI:** 10.1101/2024.10.17.24315475

**Authors:** Michael Poeschla, Uma P. Arora, Amanda Walne, Lisa J. McReynolds, Marena R. Niewisch, Neelam Giri, Logan Zeigler, Alexander Gusev, Mitchell J. Machiela, Hemanth Tummala, Sharon A. Savage, Vijay G. Sankaran

## Abstract

Variable expressivity, where individuals carrying identical genetic variants display diverse phenotypes, presents an important challenge in clinical genetics. This is exemplified by the telomere biology disorders (TBDs), which exhibit tremendous clinical heterogeneity despite their presumed monogenic nature, even among individuals harboring the same pathogenic variant. Here, we studied cohorts of patients with TBDs and population biobanks to demonstrate that common genome-wide polymorphisms associated with variation in telomere length in the general population combine with large-effect causal variants to significantly impact TBD expressivity. We go on to show that polygenic variation can contribute to expressivity within a single family with a shared large-effect causal variant, and that common and rare variation converge on a shared set of genes implicated in telomere maintenance. By elucidating the role of common genetic variation in rare disease expressivity in TBDs, these results provide a framework for understanding phenotypic variability in other presumed monogenic disorders.

## Introduction

Variable expressivity, the phenomenon where individuals with identical genetic variants display diverse clinical presentations, remains poorly understood. Deeper insights into the mechanisms underlying variable expressivity are critical for accurate genetic prognostication in clinical medicine [1,2]. While a variety of factors could contribute to variable expressivity, one proposed mechanism is common genetic variation modifying the effect of high-impact rare alleles. Supporting this, in complex diseases, polygenic variation has been shown to modify the phenotypic expression of high-impact variants [3–10]. Additionally, in presumed monogenic disease, individual loci have been identified which modify risk [11–13]. However, in extremely rare monogenic diseases, where pathogenic variants might be assumed to overwhelm any impact of common variation [14], the extent to which genome-wide polygenic variation affects expressivity has not been characterized. This in part stems from a lack of quantifiable phenotypic variability in rare disease cohorts, as well as from a paucity of individuals harboring these rare and deleterious alleles in population biobanks.

To explore the potential impact of polygenic variation on expressivity in rare monogenic disease, we focused on dyskeratosis congenita and related telomere biology disorders (TBDs), which exhibit remarkable clinical heterogeneity despite their presumed monogenic nature. The TBDs are extremely rare disorders - dyskeratosis congenita (DC), the archetypal TBD, has an estimated prevalence of approximately one in one million in the general population - caused by pathogenic germline variants in genes that regulate telomere length, maintenance, structure, and function [15–62]. Variants in these genes display striking variable expressivity related to symptom severity, age of onset, and organ involvement. For example, some carriers of known pathogenic variants in TBD-associated genes present with severe childhood-onset bone marrow failure, while others present in adulthood with idiopathic pulmonary fibrosis or liver disease, and others may have no associated clinical presentation at all (**Figure 1A)** [50,63–66]. The underlying causes of this clinical heterogeneity are poorly understood. Genetic anticipation, where disease manifestations present earlier or become more severe in successive generations, has been suggested to play a role in families with TBDs, but cannot explain all the heterogeneity present [67,68].

**Figure 1:**
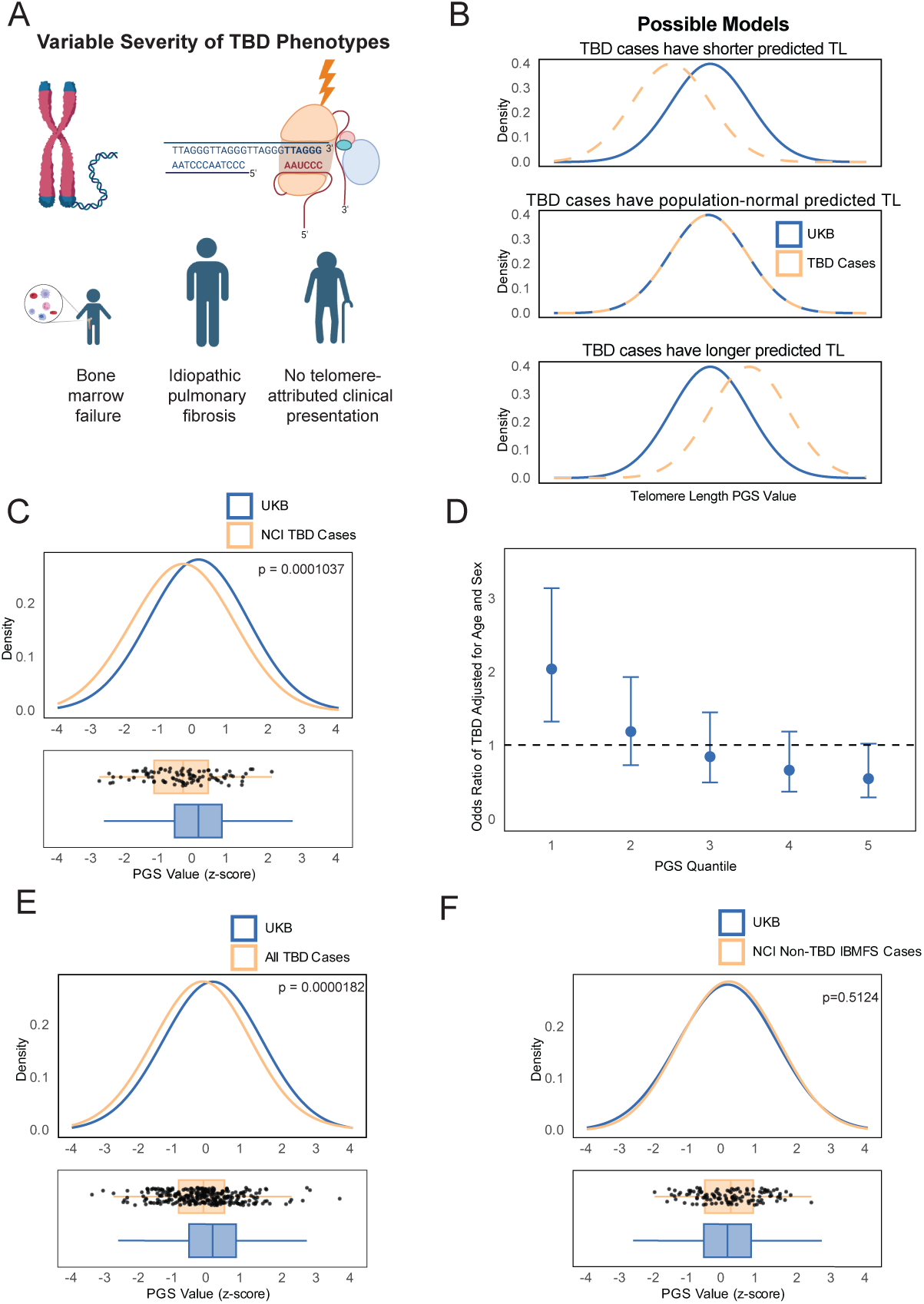
Polygenic modification of TBD expressivity in disease cohorts. A) Illustration. TBD-associated germline variants affect genes involved in telomere length and integrity. Variable expressivity of TBD variants results in diverse phenotypic presentations and age of onset. B) Schematic of distribution of TBD-case telomere length polygenic scores compared to biobank populations, under different hypotheses. If common variation affecting telomere length contributes to TBD expressivity and thus ascertainment in disease cohorts, left-shifted (towards shorter TL) PGS distribution would be expected (top panel). The null hypothesis is that TBD high-impact variants overpower any effects of common variation (central panel). An alternative hypothesis is that common variation predisposing to long telomere length protects TBD variant-carriers from severe phenotypes and mortality; under this model, conditional on surviving in order to be included in a cohort, a right-shifted PGS could be observed (bottom panel). C) Distribution of telomere length PGS in NCI TBD cases compared to the UK Biobank (Welch’s two-tailed t-test, p = 1.037E-4). D) Odds ratio of case-control status versus telomere length PGS quintile, NCI TBD cases. E) Comparison of meta-analysis of telomere length PGS distribution in NCI and DCR cases vs UK Biobank (Welch’s two-tailed t- test, p = 1.82E-5). F) Comparison of NCI non-TBD IBMFS case telomere length PGS vs UK Biobank (Welch’s two-tailed t-test, p = 0.5124).

As impaired regulation of telomere maintenance is causal for the TBDs and the underlying biological mechanisms are relatively well understood, this group of diseases presents a unique opportunity to test the idea of polygenic modification affecting variable expressivity [15,16,69,70]. Rare variants in genes that cause TBDs have been identified, and conversely, common variants that collectively underlie a considerable fraction of population-level variation in telomere length have been identified in diverse cohorts [71–74]. Therefore, we hypothesized that common genome-wide genetic polymorphisms, which contribute to the inter-individual variation in telomere length in the general population, may combine with large-effect monogenic TBD-causal genetic variants to impact variable expressivity in TBDs.

Here, we show that common genetic variation across the entire genome contributes to variable expressivity in telomere biology disorders, both in TBD cohorts enriched for individuals presenting with severe childhood-onset bone marrow failure or other hematologic manifestations, as well as in adult biobank cohorts. We further demonstrate that polygenic variation can contribute to variable disease manifestations within a single family with a shared high-impact causal variant, and that common and rare variation converge on the same biological mechanisms implicated in telomere maintenance. These results provide a framework for exploring the effects of polygenic variation on variable expressivity in rare diseases.

## Results

### Common genetic variation associated with telomere length contributes to TBD variant expressivity in inherited bone marrow failure syndrome cohorts

To assess whether common genetic variation affects expressivity in TBDs, we developed polygenic scores (PGS) using genome-wide common single-nucleotide polymorphisms associated with telomere length [71,72,75]. For a given individual, these scores provide a personalized estimate of the combined effect of common genetic variants across the genome that increase or decrease telomere length [76,77]. While these common polymorphisms underlie subtle variation in telomere length in the healthy population, we reasoned that the PGS might have a more profound impact in the context of high-impact monogenic alleles that are causal for the TBDs. We therefore applied the most predictive PGS **(Table S1)** to the National Cancer Institute (NCI) longitudinal cohort of individuals with inherited bone marrow failure syndromes, including those with TBDs (ClinicalTrials.gov Identifier: NCT00027274). The NCI cohort included 92 patients with dyskeratosis congenita and related TBDs and is significantly enriched for individuals presenting with bone marrow failure and other severe phenotypes [78].

We reasoned that if both monogenic mutations and polygenic predisposition to short telomeres contribute to the clinical severity of TBDs and the likelihood of early-onset manifestations, the distribution of genetically predicted telomere length in this clinically ascertained cohort would be shifted towards shorter telomeres compared to the population average (**Figure 1B**). Consistent with this, individuals with TBDs enriched for early-onset bone marrow failure phenotypes (n = 92) have a median polygenic score 0.43 standard deviations (SDs) shorter than the UK Biobank (p=1.04E-4), and 0.35 SDs shorter than the external All of Us cohort (p=5.82E-4) (**Figure 1C & S1B**). To estimate the effect size of the polygenic contribution, we binned the PGS distribution into quintiles based on the UK Biobank population distribution and calculated odds ratios with the number of TBD patients in each quintile representing cases and with UK Biobank participants as controls. Individuals in the lowest quintile of genetically predicted telomere length had approximately twice the odds of being a TBD case compared to those in other quintiles (**Figure 1D**). We validated these findings in a separate cohort with 190 TBD patients from the Dyskeratosis Congenita Registry (DCR) at Queen Mary University of London [79]. The DCR cohort has a broader referral base and is less enriched for reported severe bone marrow failure compared to the NCI cohort [78,79]; consistent with this, we observe a slightly attenuated, but consistent effect as our original NCI discovery cohort (median difference −0.21 SDs, p = 0.009) (**Figure S1C**). A combined analysis across both TBD cohorts further demonstrated a consistent and strongly significant association between an individual’s PGS and their odds of having a TBD (median difference −0.25 SDs, p = 1.82E-5) (**Figure 1E and S1D**).

Interestingly, in both the NCI and DCR cohorts, a subset of patients harbor no known causal high-impact TBD mutation. These patients may have lower-effect-size mutations that past TBD gene discovery efforts have been unable to detect. Under a simple liability-threshold model in which rare large-effect variants, common small effect-variants, and environmental effects combine to drive disease risk, patients with no identified large-effect variant are expected to have a more significant polygenic contribution to their disease risk on average [80,81]. To test this, we separated patients with and without a known TBD variant. While both patients with and without a known variant have a significantly shifted PGS predictive of short telomeres compared to the population average, those with no known variant had a greater PGS burden (**Figure S1E**), supporting a model in which polygenic variation contributes to TBD risk and expressivity.

We considered the possibility that population stratification and other demographic factors contributing to differences in the PGS across populations could underlie the observed differences [82–85]. To control for this, we restricted our analyses to demographically matched individuals in the disease cohorts and population biobanks (**Methods**). As a negative control, we also assessed cases of non-TBD inherited bone marrow failure syndrome cases that included individuals diagnosed with Diamond-Blackfan Anemia, Fanconi Anemia, and Shwachman-Diamond Syndrome, and that came from the same NCI bone marrow failure syndrome cohort as the TBD patients. For these conditions, telomere length does not drive the disease process, and non-telomere related gene mutations are implicated [69,70,86–88]. As expected, the polygenically predicted telomere length for these patients is indistinguishable from the UK Biobank population average (p = 0.51) (**Figure 1F**), and the telomere length PGS for the TBD patients was 0.49 standard deviations shifted towards shorter predicted telomeres compared to non-TBD inherited bone marrow failure syndrome patients (p = 3.99E-4), indicating that the observed phenomenon is telomere disease-specific and is unlikely to be due to population stratification (**Figure S1F**).

### Polygenic variation associated with telomere length impacts TBD expressivity in population biobanks

Taken together, our analyses of the NCI and DCR cohorts indicate that common genetic variants associated with short telomere length contribute to ascertainment as a TBD case in disease cohorts enriched for individuals with childhood-onset bone marrow failure. We reasoned that the reverse should also be true: TBD causal variants should be present in adult population biobanks, and adults with a pathogenic variant who avoided the severe childhood-onset manifestations of TBDs should not have polygenically predicted short telomere length (**Figure 1A**). To test this, we examined the UK Biobank for carriers of variants in the genes known to cause TBDs **(Methods)**.

To be maximally comprehensive while maintaining stringency, we defined multiple variant sets, given that each variant annotation approach has limitations for predicting true pathogenicity.[89] First, we included variants annotated in ClinVar as causing dyskeratosis congenita or a related TBD with high confidence, hereafter referred to as “ClinVar Pathogenic.” Applying quality-control, dominance and ancestry filters, we identified 213 variant carriers (**Figure S2A and S2B**). Next, we defined a more restrictive subset of ClinVar variants including only carriers of variants specifically annotated to cause childhood-onset TBDs in a dominant manner and male carriers of *DKC1*, resulting in 22 variant carriers, referred to as “ClinVar Dominant-Acting”. Finally, to be maximally inclusive of potentially pathogenic variants which may not have been annotated in ClinVar, we defined a set of predicted pathogenic rare coding variants in TBD genes using a consensus of Ensembl Variant Effect Predictor, LOFTEE, and AlphaMissense annotations, resulting in 1666 carriers in the UKB (“Consensus Predicted Pathogenic”) (**Methods**) [90–92]. Genes which harbor TBD-causal mutations are often classified based on mode of inheritance [15,16,93]. For all of these analyses, we excluded genes which cause TBDs in an exclusively autosomal recessive manner, given the challenges present in determining phase of mutations (**Methods**). Supporting the pathogenicity of the variants in the associated sets, individuals in each group in the UK Biobank had shorter measured TL compared to non-carriers. Furthermore, the magnitude of effect was concordant with the expected order of pathogenicity, with the most inclusive Consensus Predicted Pathogenic cohort associated with the smallest average decrease in TL (0.31 SDs), the more restrictive ClinVar Pathogenic cohort associated with a 0.85 SD decrease in mean TL, and the most restrictive ClinVar Dominant-Acting cohort showing the largest average decrease in TL (1.15 SDs) (**Figure 2A**).

**Figure 2:**
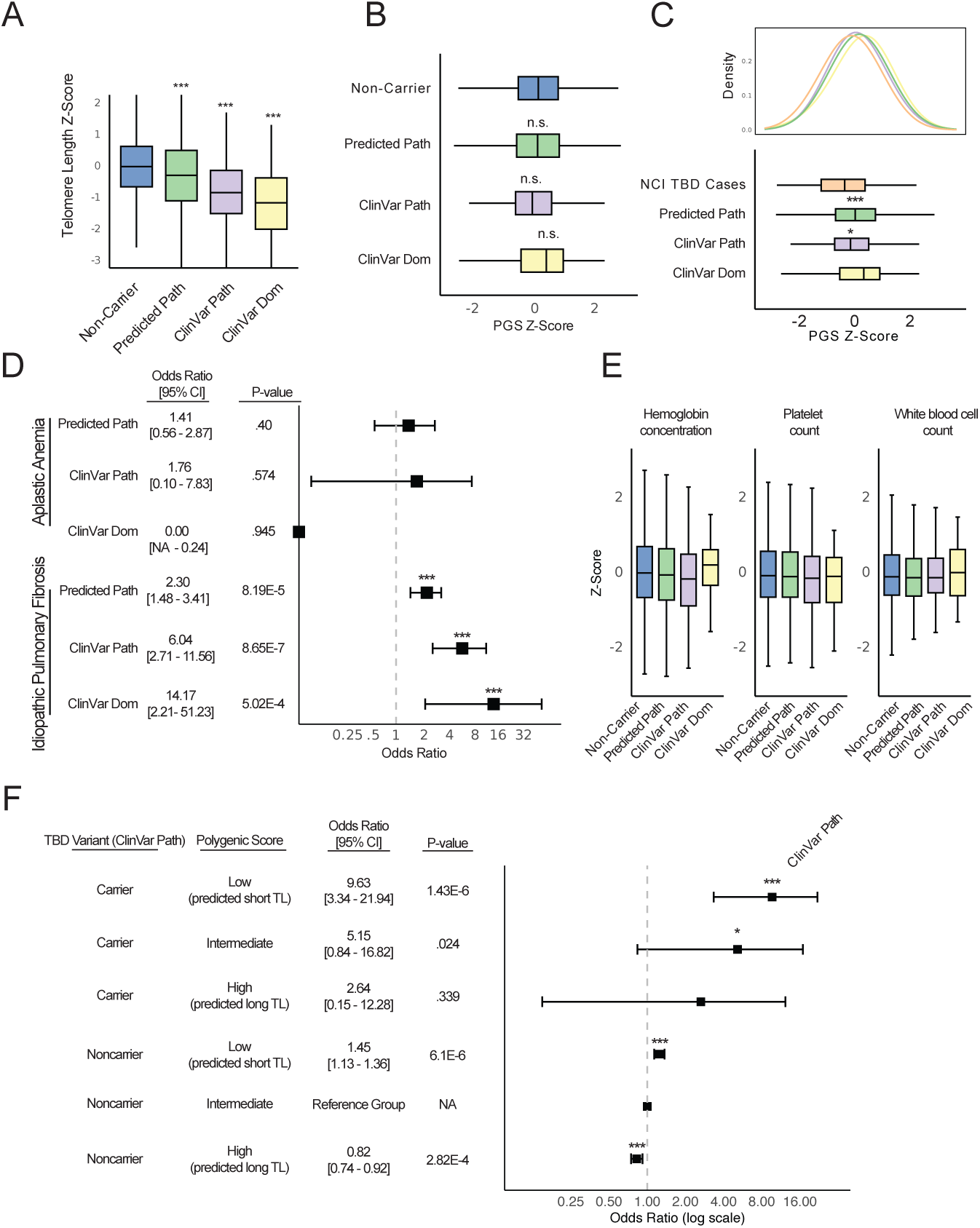
Polygenic modification of TBD expressivity in the UK Biobank. A) Measured telomere length in UK Biobank non-carriers and carriers of pathogenic TBD variants (pairwise t-tests with Bonferroni multiple testing correction, alternative = “less”: non-carrier vs Predicted Pathogenic: p = 5.01E-22; non-carrier vs ClinVar Pathogenic: p = 4.59E-23; non-carrier vs ClinVar Dominant-Acting: p = 1.86E-3) B) PGS common genetic contribution to telomere length in UK Biobank non-carriers and carriers of pathogenic TBD variants (pairwise t-test with Bonferroni multiple testing correction, alternative = “two-sided”: non-carrier vs Predicted Pathogenic: p = 1; non-carrier vs ClinVar Pathogenic: p = .20; non-carrier vs ClinVar Dominant-Acting: p = 1). C) PGS common genetic contribution to telomere length in NCI cases compared to UKB pathogenic variant carriers (pairwise t-test with Bonferroni multiple testing correction, alternative = “greater”: TBD case vs Predicted Pathogenic: p = .00087; TBD case vs ClinVar Pathogenic: p = .041; TBD case vs ClinVar Dominant-Acting: p = .090). D) Odds ratios of aplastic anemia and idiopathic pulmonary fibrosis in carriers of pathogenic TBD variants compared to non-carriers of pathogenic variants (logistic regression adjusting for age and sex). E) Blood cell counts in UK Biobank non-carriers and carriers of pathogenic TBD variants (pairwise t-test with Bonferroni multiple testing correction: no comparisons met threshold of p < 0.05). F) Odds ratios of idiopathic pulmonary fibrosis in UK Biobank stratified by PGS tertile (top third, middle third, and lowest third) and ClinVar Path variant-carrier status, using the non-carrier intermediate group as the control group (logistic regression adjusting for age, sex and first 4 ancestry PCs).

Despite having short telomeres, all three variant carrier cohorts had a population-normal polygenic contribution to telomere length on average compared to non-carriers of TBD variants in the UK Biobank (**Figure 2B**), and significantly longer predicted telomere length than the TBD cohorts (**Figure 2C**). Interestingly, the 22 carriers of mutations in the “ClinVar Dominant-Acting” set with the most severe mutations, had right-shifted polygenic score distribution relative to non-carriers, suggestive of a protective effect that could explain why carriers of these large-effect variants escaped early-life manifestations of disease and were healthy enough to participate in the UK Biobank as adults (**Figure 2B**). Together, these findings support the idea that the risk of childhood-onset severe TBD manifestations is determined not only by large-effect causal TBD gene variants, but also by common variants that impact telomere length.

We then assessed whether these pathogenic variant carriers were enriched for childhood or adult-onset TBD phenotypes relative to non-carriers in the UK Biobank. Importantly, we found no evidence for increased risk of bone marrow failure or altered blood counts in these TBD variant carriers (**Figure 2D and 2E**). However, being a carrier of a pathogenic variant was associated with greatly increased odds of presenting with idiopathic pulmonary fibrosis (**Figure 2D**). We wondered whether common variation also affects the expressivity of these adult-onset manifestations of TBDs. Strikingly, we found that both within variant carriers and in non-carriers, telomere length PGS stratifies risk of idiopathic pulmonary fibrosis (**Figure 2F and S2D**). We assessed whether there was evidence for a non-additive interaction between polygenic effects on telomere length and pathogenic variants. After accounting for independent effects, there was no significant interaction term between variant carrier status and PGS for idiopathic pulmonary fibrosis risk, supporting a model in which polygenic risk and pathogenic variants independently contribute to adult TBD manifestations, although this analysis may be limited by power (**Methods**).

In summary, we find that carriers of TBD-causing variants in the UK Biobank, in contrast to TBD cohorts, have a population-normal PGS and no enrichment for bone marrow failure, but do have increased risk of adult manifestations of TBDs. Collectively, these findings demonstrate that both pathogenic mutations and common genetic variation associated with telomere length combine to impact expressivity.

### Within-family polygenic effects on disease risk

Having observed a significant effect of common polygenic variation associated with telomere length on TBD expressivity in large disease cohorts and population biobanks, we wondered whether polygenic variation also affects expressivity within a single family, with a shared causal variant. To explore this question, we analyzed a large kindred with multiple TBD cases and a heterozygous pathogenic variant in the *TERT* gene (ClinicalTrials.gov Identifier: NCT00027274). Of the 22 family members for whom genotype data was available, there were 12 carriers of the pathogenic *TERT* variant, three of whom had clinically diagnosed telomere biology disorders (**Figure 3A**). We restricted all analyses to the 12 *TERT* mutation carriers, comparing the three clinically affected family members to the 9 unaffected family members.

**Figure 3:**
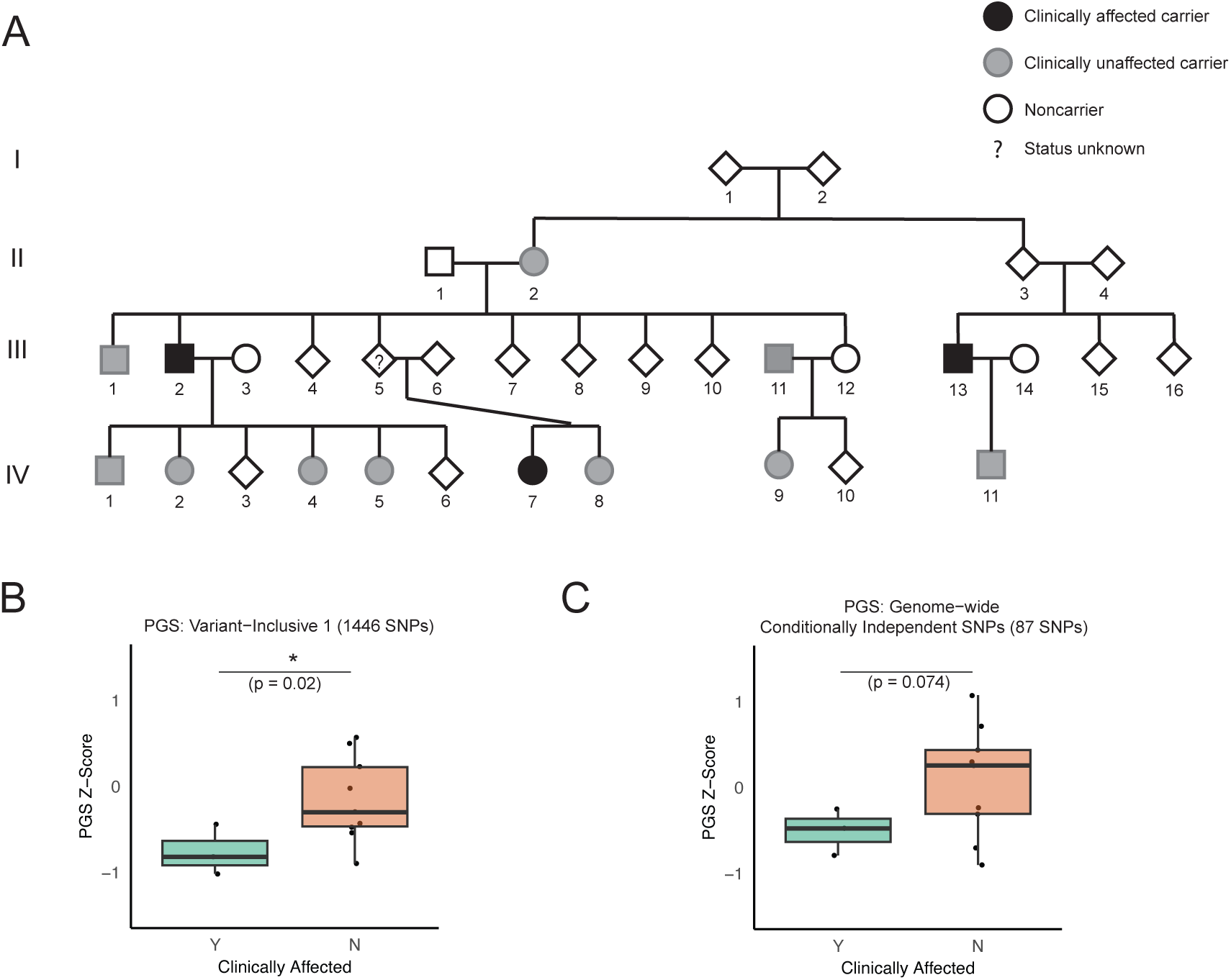
Polygenic modification of expressivity within a family. A) Pedigree depicting family with TERT variant. Black indicates case, gray indicates TERT non-case carrier, transparent indicates non-carrier. Square indicates male, circle indicates female. ? indicates unknown status, diamond indicates unknown sex. Sex information is included only for informative individuals. B) Telomere length PGS comparing cases to non-case TERT variant carriers for Variant-inclusive PGS Score 1 (linear mixed model with kinship as random effect, see **Methods**). C) Telomere length PGS comparing cases to non-case TERT variant carriers for COJO Independent Genome-Wide Significant PGS Score (linear mixed model with kinship matrix as random effect, see **Methods**).

We reasoned that the strict clumping and pruning approach that was optimal to construct polygenic scores in the UK Biobank may not be best-suited to detect relatively subtle within-family common genetic variation, given significant shared variation among the family members. Therefore, we constructed multiple polygenic predictors including more variants, reasoning that this would be more likely to pick up any subtle differences that exist within a family. As an orthogonal approach, we also constructed a polygenic score including all conditionally genome-wide significant SNP signals [94], with the idea that this would enable detection of multiple independent effects on different haplotypes which could be segregating within this family [94]. We accounted for family structure using a linear mixed model with kinship as a random effect. Remarkably, across all tested approaches, the clinically affected family members had a more negative PGS than the unaffected variant-carrying family members, indicating a greater burden of polygenic variation associated with shorter telomeres (**Figures 3B, C and S3B, C**).

Thus, even in the relatively controlled setting of a family with a shared causal variant and overall similar genetic background, we show that random segregation of common variants that alter disease biology can contribute to variable expressivity. These results suggest that common genetic variation may help to explain why some family members with a disease-causing variant have severe symptoms, while others remain clinically unaffected.

### Convergence of common and rare genetic variation in telomere biology disorders

A key question regarding polygenic modifiers of disease is whether common and rare variation converges on the same genes and biological pathways [95]. In autism spectrum disorder, examples of convergence of common and rare variation at the same loci have been observed.[96] Similarly, in Hirschsprung’s disease and craniosynostosis, shared signaling and regulatory pathways appear impacted by rare and common risk variants [6,9,11]. In contrast, in sickle cell disease and beta-thalassemia, common genetic variation largely impacts disease expressivity through modulation of fetal hemoglobin gene transcriptional regulation, a mechanism distinct from the primary disease-causing mutations that alter adult hemoglobin [10,97]. Having shown that common polygenic variation affects expressivity in telomere biology disorders, we asked whether these variants affect disease expressivity through the same or different genes as the causal high-impact variants underlying TBDs.

In the TBDs, causal variants affect genes regulating telomere length, maintenance, and function [16,93]. Using multiple gene prioritization approaches (**Methods**), we found that common variation associated with telomere length and TBD expressivity implicates genes (**Figure 4, left panel**) that strongly overlap the set of genes implicated as high-impact monogenic variants in TBDs (p = 5.41E-17) (**Figure 4, right panel**). The polygenic variants are primarily noncoding, with >97% of credible set variants mapping to introns or intergenic regions (**Figure S4B**). These variants show a striking enrichment for enhancers in CD34^+^ hematopoietic stem and progenitor cells, possibly explaining the association we observe in bone marrow failure and likely underlying impacts in these progenitors for all blood and immune cells (**Figure S4C)**. Collectively, these findings suggest a model in which common, noncoding variation converges upon the same genes implicated by high-effect Mendelian coding mutations.

**Figure 4:**
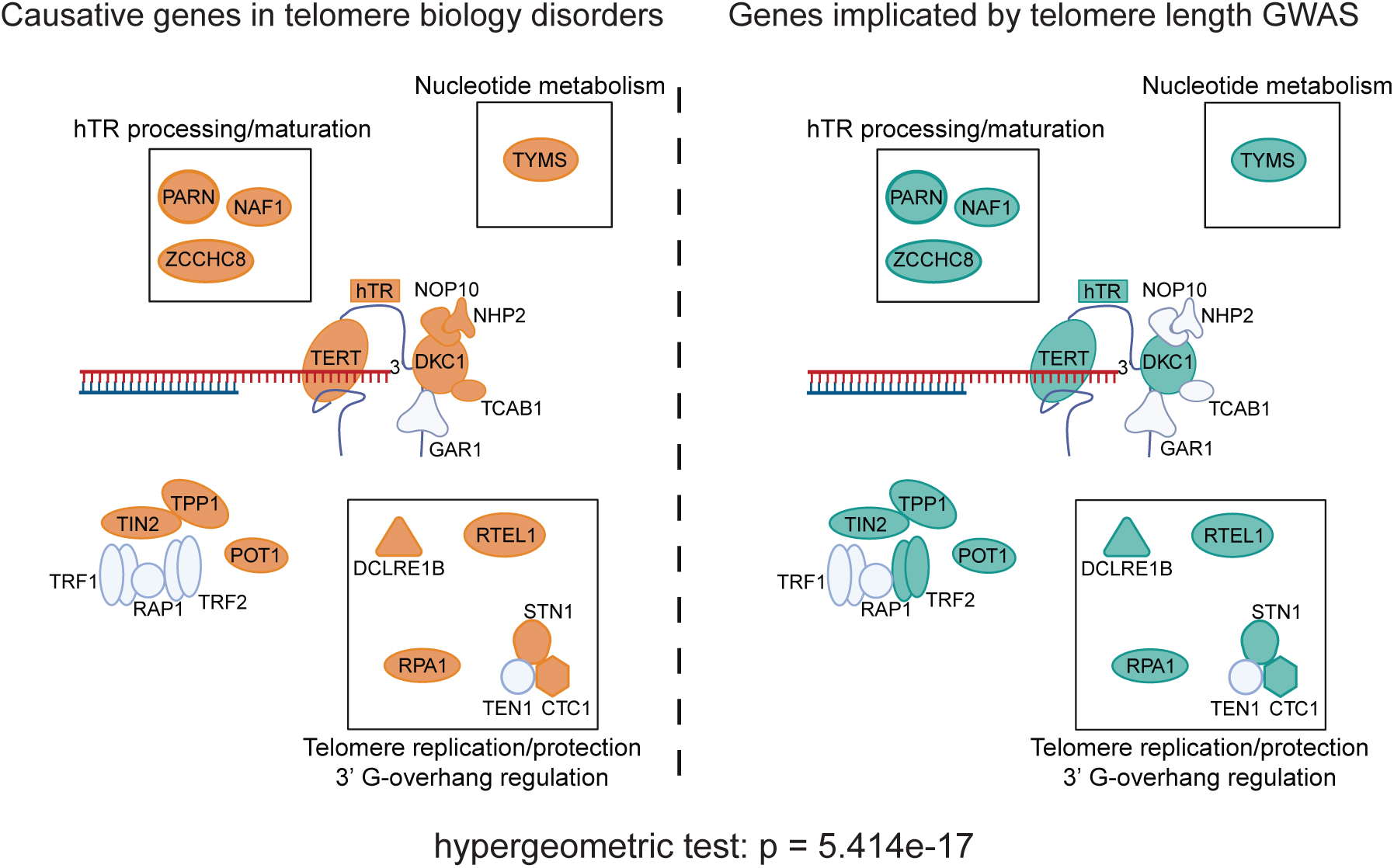
Common and rare genetic variation converges in telomere biology disorders. Genes with mutations which cause telomere biology disorders show strong overlap with genes underpinning common-variant associations with telomere length. **Left panel:** Likely causal genes associated with common SNPs included in telomere length PGS. **Right panel:** Genes implicated in telomere biology disorders (gene annotations and schematic adapted from Revy et al.[16]

## Discussion

Variable expressivity is an important challenge for understanding rare, monogenic disorders, as well as for diagnosing individuals and for determining disease prognosis [1]. Because telomere length is a measurable population-level trait, the TBDs provide a unique opportunity to test the contribution of common genome-wide variation to phenotype expressivity in a rare, presumed monogenic disease. By utilizing both disease cohorts enriched for severe, childhood-onset TBD phenotypes as well as population biobanks that are depleted of individuals with severe pediatric disease [98], we show that even in a rare-disease setting where pathogenic variants are expected to be highly penetrant, common variation plays an important role in determining phenotypic expressivity.

First, we show that common variation affecting telomere length influences the severity and clinical presentation of telomere biology disorders. We find that individuals in disease cohorts enriched for cases with severe childhood-onset TBD manifestations have a left-shifted PGS compared to the general population, indicating that polygenic variants associated with telomere length contribute to disease liability. Importantly, there is phenotypic heterogeneity in these cohorts, and we might underestimate the impact of polygenic variation on expressivity as a result. Next, we show that carriers of these same variants in the UK Biobank, a population depleted of severe childhood disease, have a population-normal PGS. Consistent with this, variant carriers in the UK Biobank do not have enrichment for severe childhood-onset TBD manifestations, but are enriched for the adult-onset TBD presentation of idiopathic pulmonary fibrosis. We also show that telomere length PGSs and rare variants independently contribute to the likelihood of having idiopathic pulmonary fibrosis in the UK Biobank. We further demonstrate that within a single family sharing a rare disease-causing variant, common genetic variation can affect an individual’s likelihood of developing the disease. Finally, we show that common, noncoding variants and rare, highly penetrant coding variants converge on genes that regulate telomere length and maintain telomere homeostasis.

Our work builds upon previous studies which have shown that polygenic variation interacts with high-impact variants in relatively more common disease settings [3,4,7,8]. Here, we have identified an example of polygenic modifiers impacting a rare disease by affecting the same genes and biological mechanisms as high-impact rare variants. Future work will likely uncover other examples, as exemplified through studies of disease modifiers such as fetal hemoglobin, where modifiers affect phenotypes through distinct biological mechanisms [10,97]. Increasingly large population sequencing studies have identified many individual rare and common variant associations with traits.[89] However, for many phenotypes, it is still challenging to explain why some variant carriers will present with severe disease, while others will have more moderate presentations, or not have any clinical presentation at all [1]. Our findings lay the groundwork for future work to precisely quantify the polygenic contribution to variable expressivity across a range of common and rare diseases.

Our measurement of common variant effects on telomere length are derived from qPCR-based measurement in white blood cells. Mean telomere length is only a correlated proxy for the length of the shortest telomere, which is likely the more biologically meaningful measurement in telomere homeostasis [99,100]. Furthermore, telomere length is correlated yet variable across cell and tissue types, and DC is a multisystem disease. Thus, while mean leukocyte telomere length is the best available metric at a population level to assess common genome-wide variation impacting telomere homeostasis, this measurement provides an imperfect estimate of the common genetic variant effects which contribute to disease expressivity in the TBDs, and better telomere measurements that are emerging, applied across multiple tissues, will enable more precise population-level analyses in the future [101,102]. Furthermore, both disease cohorts and population biobanks have strengths and weaknesses for assessing pathogenic variant penetrance and expressivity. Disease cohorts may overestimate variant pathogenicity, and adult biobanks are likely to underestimate variant impact [103]. For this reason, in this study we utilize both disease cohorts and population biobanks, which reveal complementary insights.

Finally, TBDs are heterogeneous, and mode of inheritance as well as specific causal variants have been shown to be associated with variation in outcome [15]. In this study, we grouped different genotypes together to best power analyses of the effects of common genetic variation, likely obscuring some effect heterogeneity. Future, larger studies will be necessary to identify variant- and gene-specific interactions affecting variable expressivity.

## Data Availability

The telomere length summary statistics will be deposited at the NHGRI-EBI GWAS catalog upon publication. All code and accompanying de-identified data necessary to reproduce all plots and analyses presented in this study will be deposited at GitHub upon publication. Any additional de-identified data generated in this study are available upon reasonable request to the Lead Contact, provided the request is consistent with study consent documents.

## Acknowledgments

We thank members of the Sankaran laboratory and colleagues including D. Tang, M. Talukdar, T. de Lange, D. Nathan, and D. Ginsburg for valuable discussions on this work. This work was supported by National Institutes of Health grants R01DK103794, R01HL146500, R01CA265726, R01CA292941 (to V.G.S.), the New York Stem Cell Foundation (to V.G.S.), the Mathers Foundation (to V.G.S.), a gift in memory of Jan Ellen Paradise, MD to Boston Children’s Hospital (to V.G.S.), and the Howard Hughes Medical Institute (to V.G.S.). This research has been conducted using the UK Biobank Resource under application number 31063. Figure schematics were prepared with BioRender. M.P. was supported by National Institutes of Health grants T32GM007753 and T32GM144273. The work of S.A.S., L.Z., M.R.N, N.G., and L.J.M was supported by the intramural research program of the Division of Cancer Epidemiology and Genetics, National Cancer Institute. The content is solely the responsibility of the authors and does not necessarily represent the official views of the National Institute of General Medical Sciences or the National Institutes of Health. V.G.S. is an Investigator of the Howard Hughes Medical Institute.

## Author contributions

M.P. and V.G.S. conceived and designed the study. M.P. and U.P.A. performed computational and statistical analyses. A.W., L.J.M., N.G., L.Z., H.T., and S.A.S. provided genotype and phenotype data from disease cohorts. A.W., L.J.M., M.R.N., N.G., A.G., M.J.M., H.T., and S.A.S. contributed ideas and insights on the analyses. M.P. and V.G.S. wrote the manuscript with input from all authors. V.G.S. supervised all analytical aspects of this work. All authors read and approved the final version of the manuscript.

---

**Supplementary Figure S1:**
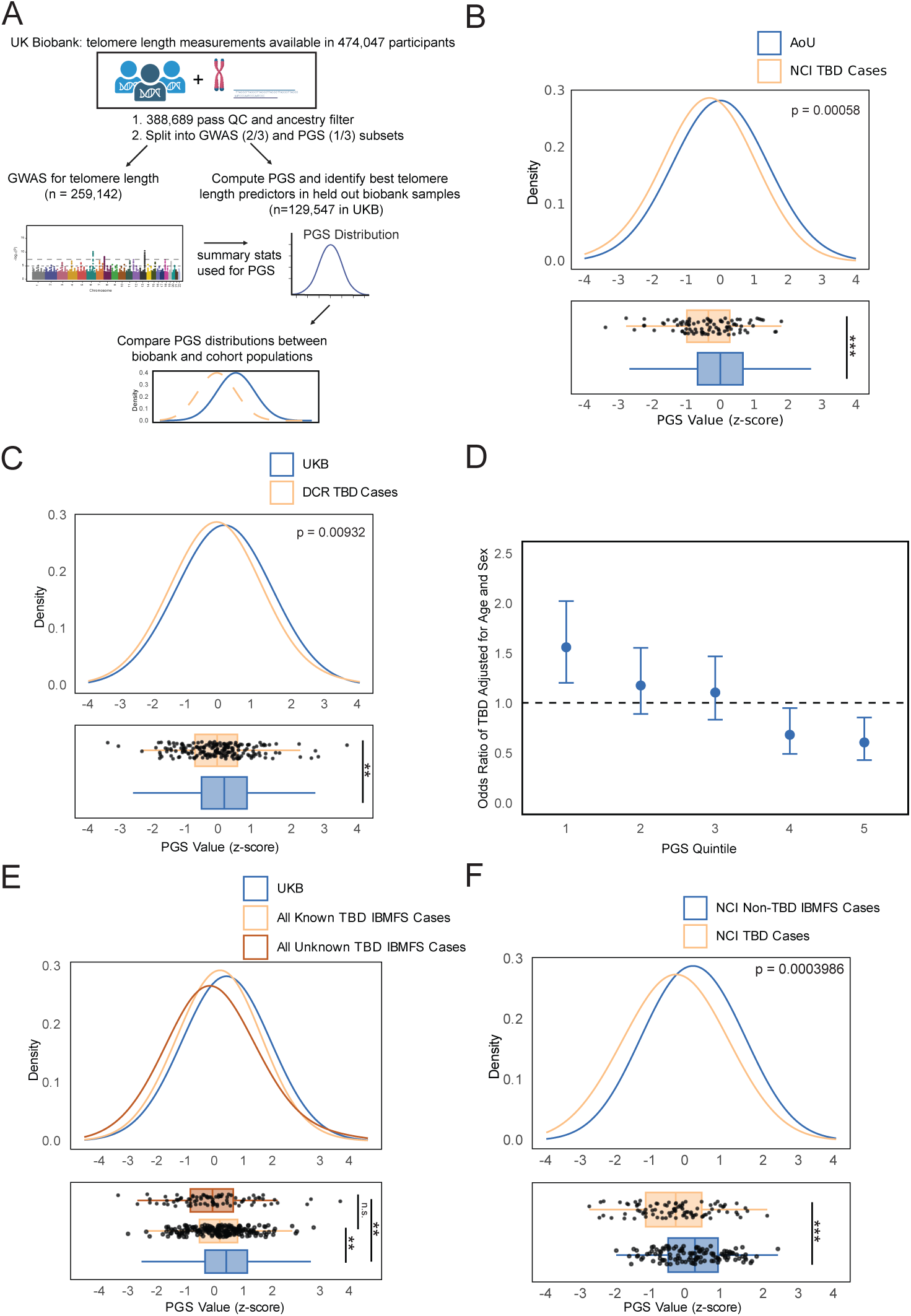
Design and analysis of TBD disease cohorts. A) Schematic of study design to assess polygenic variation affecting telomere length across biobank and cohort populations. B) Distribution of telomere length PGS in All of Us cohort compared to the UK Biobank (Welch’s two-tailed t-test, p = 5.8E-3). C) Distribution of telomere length PGS in DCR cases compared to the UK Biobank (Welch’s two-tailed t-test, p = 9.32E-3). D) Odds ratio of case-control status versus telomere length PGS quintile, meta-analysis of NCI and DCR cases vs UK Biobank. E) Comparison of known and unknown TBD-causing mutations for all meta-analyzed cases and UK Biobank (pairwise Welch’s two-tailed t-test: UKB vs known IBMFS cases: p = 1.393E-3; UKB vs unknown IBMFS cases: p = 2.788E-3; known vs unknown IBMFS cases: p = 0.2157). F) Comparison of NCI TBD cases and NCI non-TBD inherited bone marrow failure syndrome case telomere length PGS (Welch’s two-tailed t-test, p = 3.986E-4).

**Supplementary Figure S2:**
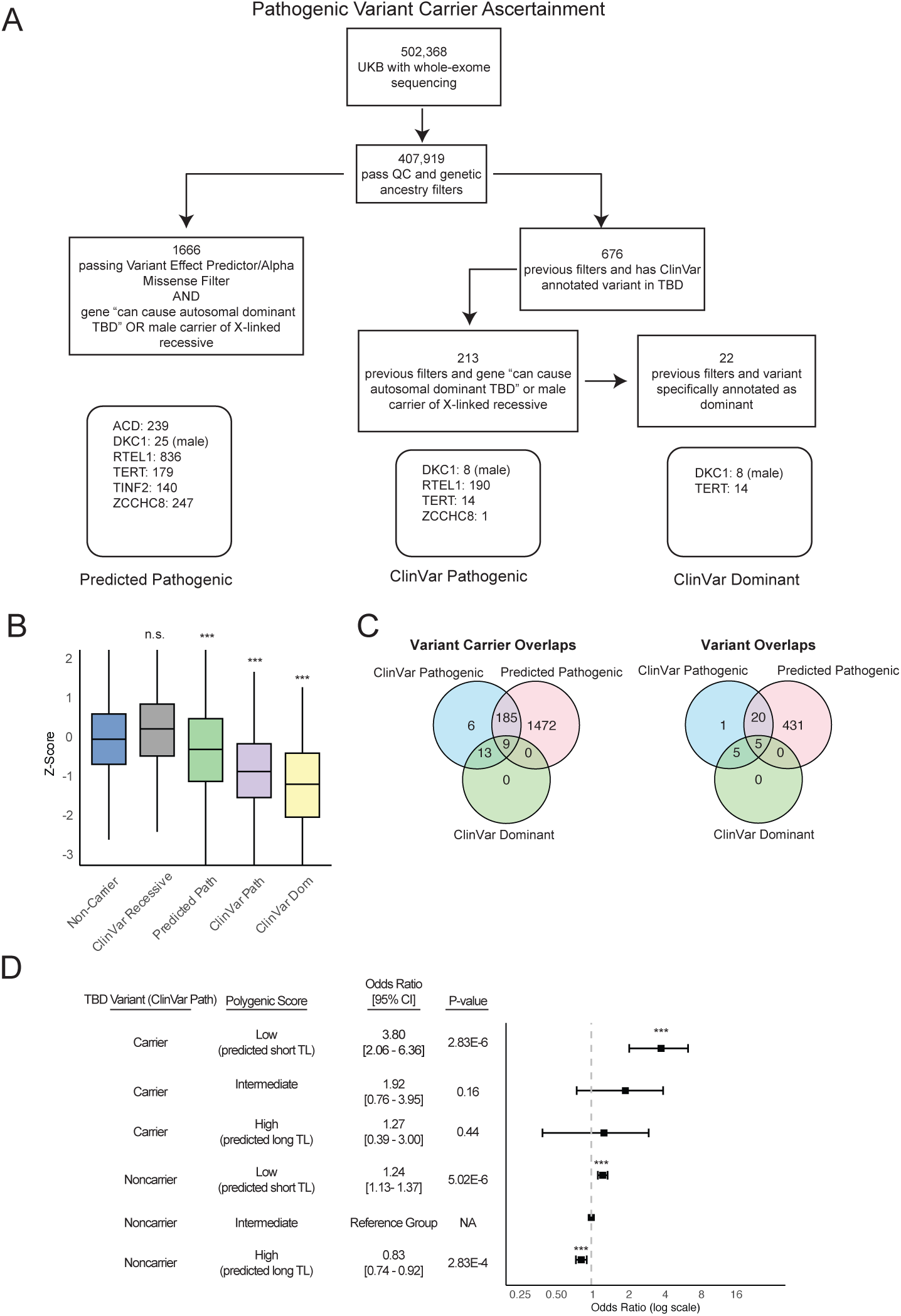
Ascertainment strategy and analysis of UK Biobank. A) Variant carrier ascertainment strategy in UK Biobank, and summary of number of mutation carriers for each gene in three variant carrier classes. B) Measured telomere length in UK Biobank non-carriers and carriers of pathogenic TBD variants, showing carriers of ClinVar mutations in recessive genes (no enrichment for short telomere length) (pairwise t-test with Bonferroni multiple testing correction, alternative = “less”: non-carrier vs Predicted Pathogenic: p =9.08e-22; non-carrier vs ClinVar Pathogenic: p =2.28e-23; non-carrier vs ClinVar Dominant: p = 9.31e-4; non-carrier vs ClinVar Recessive: p = 1). C) Number within and overlaps between each pathogenic category for variant carriers and individual variants. D) Odds ratios of idiopathic pulmonary fibrosis in UK Biobank stratified by PGS tertile (top third, middle third, and lowest third) and Predicted Pathogenic variant-carrier status, using the non-carrier intermediate group as the control group.

**Supplementary Figure S3:**
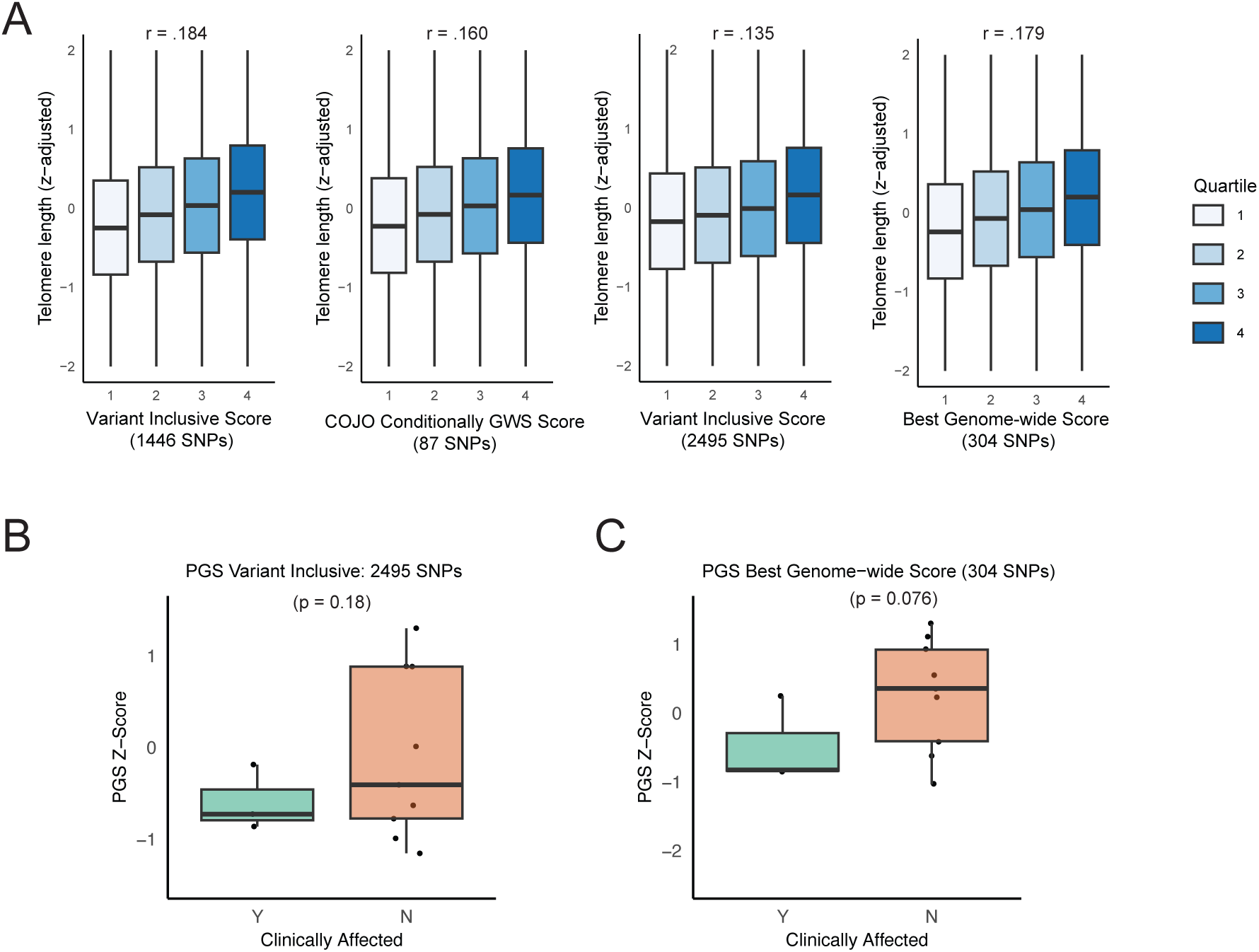
Pedigree data strategy and analysis. A) Measured telomere length by PGS quartile across different PGS scores constructed for pedigree SNPs (Spearman correlation between quartile and adjusted telomere length). B) Telomere length PGS comparing cases to non-case TERT variant carriers for Variant-inclusive PGS Score 2 (linear mixed model with kinship matrix as random effect, see **Methods**). C) Telomere length PGS comparing cases to non-case TERT variant carriers using same parameters as best genome-wide SNP score (linear mixed model with kinship matrix as random effect, see **Methods**).

**Supplementary Figure S4:**
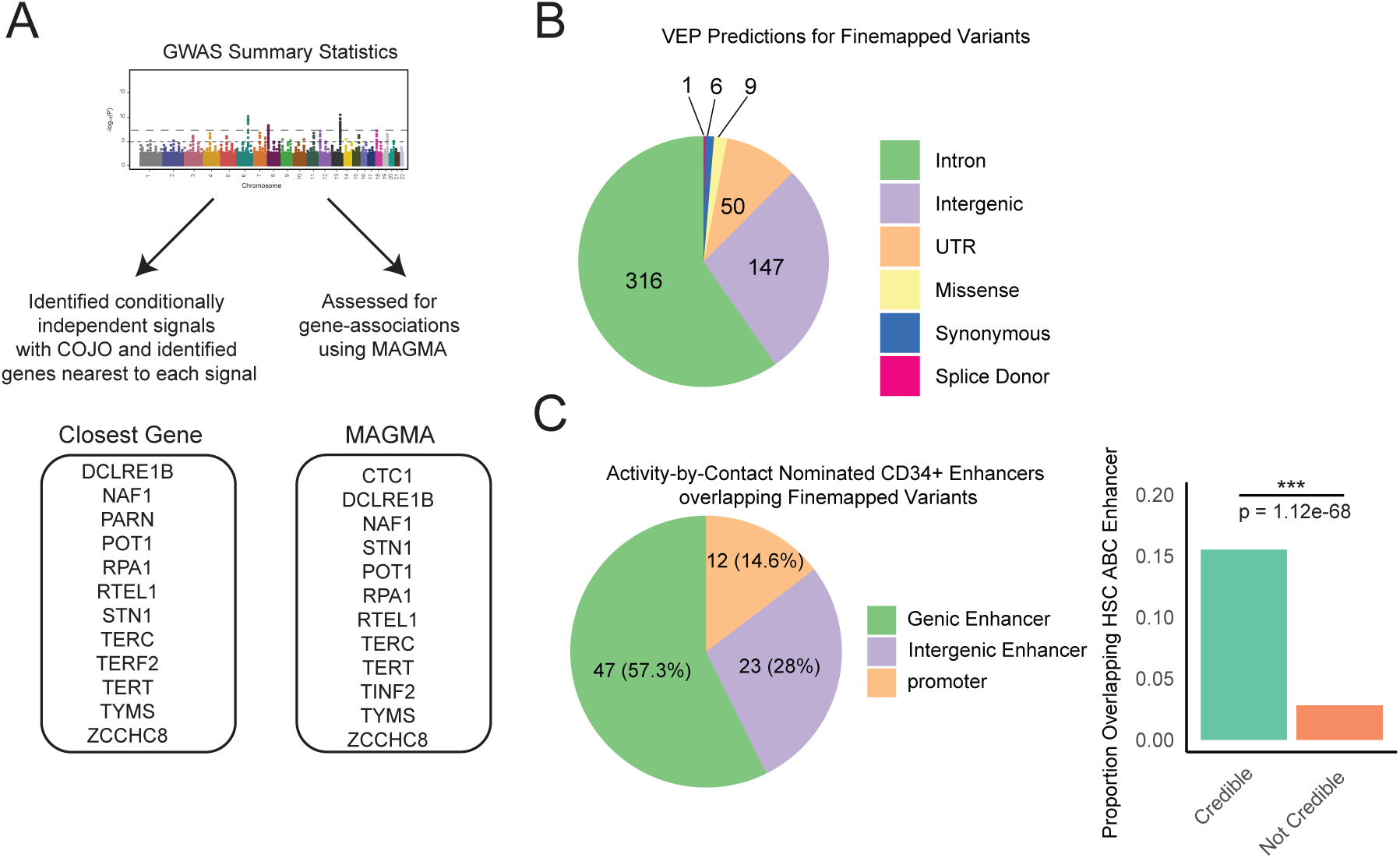
Causal gene prioritization and variant functional annotation. A) Strategy to prioritize likely causal genes. Genes closest to conditionally independent genome-wide significant signals were identified. MAGMA was used to identify genes associated with summary statistics. B) VEP predictions for fine-mapped 90% credible-set variants. C) Finemapped variants overlapping ABC enhancers in hematopoietic stem cells (enrichment computed using chi-squared test)

## Methods

### Genotype data processing from various cohorts

#### UK Biobank

The UK Biobank is a large prospective cohort with extensive phenotype and molecular data available, including whole-genome sequencing [104,105]. The UK Biobank was accessed under application number 31063. Cohort analysis details are under METHOD DETAILS - *UK Biobank*.

#### All of Us

All of Us is a longitudinal cohort that currently contains short-read whole genome sequencing data and phenotypic data from 245,394 participants. Whole genome sequencing was performed on participants to a mean depth of 30x. Informed consent for all participants is conducted in person or through an eConsent platform that includes primary consent, HIPAA Authorization for Research use of EHRs and other external health data, and Consent for Return of Genomic Results. The protocol was reviewed by the Institutional Review Board (IRB) of the All of Us Research Program. The All of Us IRB follows the regulations and guidance of the NIH Office for Human Research Protections for all studies, ensuring that the rights and welfare of research participants are overseen and protected uniformly. Cohort analysis details are under METHOD DETAILS - *All of Us*.

#### National Cancer Institute Inherited Bone Marrow Failure Syndrome samples

Processed genotype array files were obtained as part of the NCI Inherited Bone Marrow Failure Study (ClinicalTrials.gov Identifier: NCT00027274). Genotype data was available for 92 dyskeratosis congenita cases, 49 Diamond-Blackfan Anemia cases, 45 Fanconi Anemia cases, and 19 Shwachman-Diamond Syndrome cases. Genotype data was also available for 22 members of a family with a shared *TERT* mutation that included 4 dyskeratosis congenita cases. Genotype data was also available for 120 unaffected family members of dyskeratosis congenita cases in the cohort.

#### Dyskeratosis Congenita Registry Queen Mary University of London samples

Two sample sources were used as part of the DCR cohort. The first included 49 samples of unknown mutation that had previously undergone sequencing. For the second, DNA from 132 cases of known genotype was sequenced using low-pass whole-genome sequencing (mean coverage 0.5X) by Azenta Life Sciences.

#### UK Biobank

We excluded individuals with mismatches between self-reported and genetically inferred sex, possible sex chromosome aneuploidy, outliers for heterozygosity or missingness, excessive kinship, and individuals who withdrew consent at the time of analysis. Because telomere length has been reported to vary significantly across ancestry groups and our patient population is predominantly white European ancestry, we excluded individuals who were not in the white British ancestry subset defined both by self-report and by genetic ancestry [106].

#### All of Us

Only samples with matching reported sex and genetically inferred sex were included. Site level QC metrics included QUAL > 60, ExcessHet < 54.69, GQ > 20, DP > 10, AB > 0.2 for heterozygotes. Variants with population-specific allele frequency (AF) > 1% or population-specific allele count (AC) > 100, in any computed ancestry subpopulations were selected. Sites with more than 100 alternate alleles were excluded. A high-quality set of SNPs were used to determine ancestry labels for each sample based on the same labels as gnomAD and were used to filter for European (EUR) population individuals.

#### National Cancer Institute Inherited Bone Marrow Failure Syndrome samples

See experimental model and subject details. Genotype array data was obtained in PLINK file format and imputed using the TOPMed Imputation Server [107–109]. The data from the kindred with a shared *TERT* mutation was considered separately from the other TBD cases and only in pedigree-specific analyses.

#### Queen Mary University of London samples

See experimental model and subject details. For the samples of unknown genotype, processed VCF files were imputed using the TOPMed Imputation Server. The resulting FASTQ files from the Azenta sequenced samples were adapter trimmed using cutadapt and then aligned to the hg38 reference genome using bwa-mem. Since the TOPMed Imputation Server is not designed to impute from low-pass sequencing samples, these samples were imputed with GLIMPSE2 using the 1000 Genomes phase 3 release as a reference set [110,111].

### Genome-wide association analysis for telomere length

#### Phenotype and sample construction

For the telomere phenotype, we used the qPCR-based leukocyte telomere length measurements generated, batch-corrected and quality controlled in 474,074 UKB participants (UKB Data Field 22191) [72]. The phenotype was log-normalized and z-scored as in Codd et al. [71]. Of the remaining individuals, we then removed the individuals with the highest and lowest 0.5% of telomere length values to minimize the impact of outliers.

The set of filtered, phenotyped individuals was randomly split into two non-overlapping subsets. Two-thirds of the individuals (n = 259,142) were used in the GWAS analysis of telomere length, and the remaining one-third of samples (n = 129,547) were used for subsequent polygenic risk score construction.

#### Genome-wide association study for telomere length

REGENIE [112] was used to conduct a GWAS for telomere length. For Step 1 of REGENIE, the UKB genotype array files were quality controlled following the recommendations of the REGENIE developers, removing SNPs with minor allele frequency below 1%, minor allele count below 100, genotype missingness greater than 10%, sample missingness greater than 10%, and Hardy-Weinberg equilibrium test p-value exceeding 1e-15.

For Step 2 of REGENIE, we used common SNPs from the November 2023 UK Biobank whole-genome sequencing release (UKB field: 24053). The DRAGEN individual-level variant call files were filtered for minor allele frequency of at least 0.1% using BCFtools and the following quality control criteria: HWE>10e-30, INFO/AN > 0.9*AN, ExcHet >= 0.5 && ExcHet <= 1.5, FILTER=“PASS” && MAF>0.001, then merged into PLINK files for REGENIE.

Step 1 of REGENIE was run using recommended parameters with --bsize 1000. Age, sex, and the first ten genetic principal components were included as covariates. Step 2 was run using the .loco and .pred.list files output from step 1 with --quant_traits=true, and age, sex, and the first ten genetic principal components as covariates.

The summary stats were then used with one further processing step: because the qPCR-based telomere length measurement relies on HBB gene amplification as a control, the measurement results in a previously reported artifactual enrichment of genetic associations around the HBB gene [71]. Following the procedure previously employed by *Codd et al.*, we removed 1 million bases flanking either end of the HBB gene from the summary stats files before constructing polygenic risk scores.

### Polygenic risk score construction using telomere length summary statistics

#### Primary polygenic risk score construction

The summary statistics and processed whole-genome sequence variant call files described above were then used to construct polygenic risk scores using the 129,547 samples that were not included in the GWAS. As the goal of constructing these scores was for use as an instrument to assess the burden of common genetic variation affecting telomere length across different cohorts, we restricted analysis to SNPs that were well imputed (INFO > 0.6) across each of the different cohorts that were tested. This retained 5,279,945 SNPs genome-wide.

Polygenic scores were constructed using the common clumping and thresholding approach. In clumping and thresholding, index SNPs are selected, and SNPs within a prespecified distance window and above a prespecified correlation threshold are identified (“clumped”) and removed (“pruned”), to remove redundant effects from SNPs on the same haplotype, and this process is iteratively repeated across the genome. For the variants remaining after pruning, a range of p-value thresholds is then applied, removing all variants above the p-value threshold. The resulting variant sets are then used to compute polygenic scores defined as the sum of variant allele counts weighted by effect size estimates from the telomere length GWAS.

To identify the most predictive overall scores in the UK Biobank population, a grid search over linkage disequilibrium thresholds including r^2 values of [0.001, 0.005, 0.01, 0.02, 0.05, 0.1, 0.2, 0.5, 0.8] and clumping windows including [100, 250, 500, 750, 1000, 1500, 2000, 2500, 3000, 4000, 5000, and 7500] kilobases was performed. For each of these, a range of p-value thresholds was tested using PRSice-2 from 5e-8 to 5e-4 in increments of 1e-7. The most predictive score by variance explained was identified. The following flags were used for PRSice-2: --base-info INFO: 0.6, --beta, --binary-target F, --cov-col “@pc[1–10],sex,age”, --model add, using the 5,279,945 SNP set previously defined, and with z-scored log-normalized adjusted telomere length from Codd et al. as the phenotype [72,75].

From the grid search analysis, there was a range of parameter values that provided similar, greatest prediction accuracy (r^2 0.02, with LD window size 2000 and greater). Of these, the lowest parameter value was selected at which prediction accuracy was observed to plateau (r^2 0.02, with LD window size 2000) for downstream analyses, yielding a polygenic score with 304 genome-wide SNPs (**Table S1)**. The variance explained for telomere length of the best polygenic score using these parameters was 0.071 (**Table S1**).

#### Comparing polygenic risk scores across cohorts

The SNPs which were included in the best PGS score computed in the UK biobank were used to directly compute scores in each different cohort using PRCise-2 with the following parameters used to avoid following the clumping and thresholding algorithm to just compute scores directly from those SNPs: --no-clump --no-full --no-regress --fastscore. The polygenic score values were then z-scored using the UK Biobank score distribution as a population reference to enable easy interpretation. Thus, the UK Biobank score has a mean of 0 and standard deviation of 1, and all other cohort scores can be interpreted from that reference.

#### All of Us polygenic risk score construction and testing

To compare telomere length PGS between the All of Us [113] cohort and patient samples, an identical construction procedure was followed as in UK Biobank, but restricting to SNPs genotyped with high quality in All of Us, resulting in inclusion of 6,219366 SNPs for score construction. (see “Genotype data from various cohorts”). We then re-constructed PGS in the same subset of the UK Biobank as previously with these SNPs, the GWAS summary statistics described above, and the UKB measurement for telomere length used previously. We used the exact same parameters that resulted in the most predictive scores in the UK Biobank in our general analysis (r^2 0.02, with LD window size 2000). This resulted in a 253 SNP PGS (**Table S1**). We then used this score to compute PGS for telomere length in the European (EUR) population in AoU, and in the patients from the NCI cohort. As above these scores were directly computed using PRCise-2 with the following parameters used to avoid following the clumping and thresholding algorithm, to just compute scores directly from those SNPs: --no-clump --no-full -- no-regress --fastscore. The polygenic score values were then normalized to the AoU values. Thus, the AoU score has a mean of 0 and standard deviation of 1, and the cohort scores can be interpreted from that reference.

#### Plotting and comparing score distributions

Score distributions across groups were pairwise compared using Welch’s t-test with a two-sided alternate hypothesis. Distributions were plotted with kernel density estimates plotted over boxplots using ggplot2. For KDE plots, bandwidth 1 was used as a parameter.

#### Computing odds ratios

To assess the relationship between PGS and the outcome of dyskeratosis congenita, we defined dyskeratosis congenita affected individuals as cases and UK Biobank participants as controls. The PGS population values were binned into 5 equal-sized quintiles, and each observation for both cases and controls were placed in the appropriate quintile. We then calculated odds ratios and 95% confidence intervals for each quintile. For each quintile, a 2×2 contingency table was constructed, comparing the number of outcome cases within the quintile to those outside it. Fisher’s exact test was applied to estimate the odds ratio for each quintile. The standard error of the logarithm of the odds ratio was used to compute the 95% confidence intervals.

### UK Biobank pathogenic variant carrier analysis

#### Defining variant sets

For all analysis of variant carriers in the UK Biobank, the following genes weres used as a TBD gene set, defined based on genes with strong evidence of being capable of causing dyskeratosis congenita, Revesz syndrome, Hoyeraal-Hreidarsson syndrome, or Coats plus syndrome, and having a mode of inheritance reported in the literature as causing dyskeratosis congenita/other syndromic TBDs or bone marrow failure in a monoallelic or “monoallelic or biallelic” fashion, excluding genes for which only biallelic inheritance has been reported **(Table S2)** [16]. This resulted in inclusion of the following genes: *TERC, TINF2, ZCCHC8, ACD, RTEL1, TERT,* and *DKC1.* For *DKC1*, only male carriers (X-linked recessive) were included. Noncarriers were defined as any participants passing QC that did not carry one of the pathogenic variants included in the final analyses.

ClinVar variants were defined using the following query of the ClinVar database (as of July 11th, 2024):

- ((((dyskeratosis congenita) OR Coats plus) OR hoyeraal-hreidarsson) OR Revesz syndrome) AND (((“clinsig likely pathogenic”[Properties] or “clinsig likely risk allele”[Properties]) OR (“clinsig pathogenic”[Properties] or “clinsig pathogenic low penetrance”[Properties] or “clinsig established risk allele”[Properties]))) with “Review status” “At least one star” and “Classification type” “Germline”, yielding 592 variants.

The Predicted Pathogenic dataset was produced starting from all variants in the whole exome dataset that passed QC (above) with less than 1% frequency, within the start and end positions of known TBD genes (NCBI). Variants were then annotated using the Ensembl Variant Effect Predictor. The annotations were then filtered to retain all predicted “HIGH IMPACT” variants. Variants with predicted “MODERATE IMPACT” were further annotated using the AlphaMissense released pathogenicity scores, and variants predicted as “likely_pathogenic” were retained [90,91].

#### Pre-processing the UK Biobank whole-exome sequencing data

The UKB exome sequencing final release of 454,787 participants [114]. PLINK fileset was filtered for rare variants with allele frequency less than or equal to 1 percent, that passed the recommended variant-level filter requiring that at least 90% of all genotypes for a given variant - independent of variant allele zygosity - are nonmissing and have a read depth of at least 10 (i.e. DP>=10). Variants were further processed to exclude samples with missingness greater than 10%. Indels and SNVs were then extracted by filtering the processed exome files by matching on chromosome, position, reference and alternate alleles, for the variant sets defined above.

#### Identification and phenotypic analysis of carriers of whole-exome variants

We used the UK Biobank Research Analysis Platform to extract phenotype data. We first filtered individuals using the criteria described above for quality control and White British ancestry (see Methods: *UK Biobank*). We extracted phenotypic information on blood cell counts, clinical records, and telomere length. To assess for blood cell count differences, the Haemoglobin_concentration, Platelet_count, and White_blood_cell_count phenotype fields were utilized and z-scored. To assess for idiopathic pulmonary fibrosis, we used ICD-10 code of J84.1 (“other interstitial pulmonary diseases with fibrosis”). To assess for aplastic anemia, we used ICD-10 code of D61 (“Other aplastic anaemias,” which includes “Aplastic anaemia, unspecified,” “constitutional aplastic anaemia”, “idiopathic aplastic anaemia”, “other specified aplastic anaemias” and “aplastic anaemia, unspecified”).

#### Statistical analyses

For the comparisons between non-carriers and the different pathogenic carrier sets and for dyskeratosis congenita cases compared to the pathogenic carrier sets, because statistical comparisons were directly between the reference set and the pathogenic sets, pairwise t-tests were used with Bonferroni multiple testing correction. Logistic regression models were fitted to estimate the association between genetic annotations and idiopathic pulmonary fibrosis or aplastic anemia. The models included age, sex, and the first 4 principal components of ancestry as covariates. Odds ratios (ORs) with 95% confidence intervals (CIs) were extracted from each model with pathogenic carrier status as the predictor. Models were fit using the glm function in R with a binomial family, and ORs were obtained by exponentiating the model coefficients.

To test for a PGS contribution to IPF risk in variant-carriers and non-variant carriers with an interpretable odds ratio output, we created variables indicating PGS tertile and carrier status for ClinVar Path variants or Consensus Predicted Path variants (Group 1 = PGS high and carrier, Group 2 = PGS high and non-carrier, Group 3 = PGS intermediate and carrier, etc.). For each group, UKB participants were coded as 1 if part of this group, as 0 if not a member of the other pathogenic variant groups, and as NA if included in the other variant carrier groups. We then performed logistic regression adjusting for age, sex and the first 4 PCs of ancestry, with IPF as the dependent variable.

To test for an interaction between the PGS and rare variant status, we performed a similar logistic regression with age, sex, the first 4 PCs, the individual PGS values (non-binned), and ClinVar variant carrier status, and an interaction term between PGS and variant carrier status.

### Analysis of expressivity within a family

#### Pedigree analysis polygenic risk score construction

We utilized the same set of 5,279,945 SNPs to compute polygenic risk scores in the pedigree cohort. We constructed four different polygenic scores. Three were constructed taking the best score using clumping and thresholding with various sets of hyperparameters. For all scores, the LD r^2 threshold and window size for calculating LD was set, and then scores of increasing p-value were tested (starting from 5E-8, and increasing at intervals of 5E-7), and the score with the best explanatory power in the UK Biobank was selected. The following sets of clumping and thresholding hyperparameters were used: “PGS: Variant Inclusive 1”, we used an LD r^2 threshold of 0.2 and an LD window size of 2000 kb, resulting in inclusion of 1446 SNPs and then calculated the best score (**Table S1**). For “PGS: Variant Inclusive 2”, we used an LD r^2 threshold of 0.5 and an LD window size of 500 kb, resulting in inclusion of 2495 SNPs (**Table S1**). For the PGS: Best Genome-wide Conditionally Significant SNP Score, we included all conditionally significant independent non-ambiguous signals using GCTA-COJO (see Methods: *Finemapping*), resulting in inclusion of 87 SNPs (**Table S1**). Finally, we used the same parameters as our best genome-wide score, with an LD r^2 threshold of 0.02, and an LD window size of 2000 kb, including 304 SNPs as described previously. Each pedigree score was validated by binning the PGS into four quartiles and computing the Spearman correlation between the quartile and measured telomere length in the UK Biobank.

#### Family data statistical analysis

Because of high relatedness within the pedigree, a linear mixed model with a kinship matrix as a random effect was used. The kinship matrix was generated using PLINK (-- make-rel square) using the pedigree genotypes to estimate relatedness. The LMM was performed using lmm.aireml, with the PGS and an intercept term as fixed effects and the kinship matrix as a random effect. Fixed effects estimates and standard errors were extracted from the LMM results. Wald test statistics were computed as the ratio of the fixed effects to their standard errors, and p-values were derived from the Wald test statistics.

### Analysis of convergence of common and rare variation

#### Gene prioritization

From the telomere length GWAS summary statistics, genes were prioritized using two approaches. MAGMA [115] was used with parameters --gene-model snp-wise=mean. A false-discovery rate of 0.01 was applied to identify MAGMA prioritized genes. For closest gene analysis, the COJO conditionally genome-wide significant SNPs were used and the three nearest genes were identified based on location (overlapping or nearest distance to start or end) using BEDtools [116]. The MAGMA and closest gene prioritized sets were combined into a single common-variation prioritized set **(Figure S4A)**. A hypergeometric test was used to test for enrichment of the known TBD causal genes in this set.

#### Conditional and joint analysis using summary data

Conditionally independent SNP signals were identified using GCTA-COJO [117] with the telomere length summary statistics, using the UK Biobank dataset WGS PLINK fileset as a reference with the following parameters: --cojo-wind 10000 --cojo-slct --cojo-p 5e-8.

#### Finemapping

Finemapping was performed using FINEMAP [118]. Sentinel SNPs for finemapping were selected by using COJO outputs and iteratively selecting COJO independent signals with a p-value less than 5e-6, and then iteratively expanding the finemapping region if other COJO signals were found within one megabase of each signal. For each sentinel region, all SNPs were extracted from the UKB WGS PLINK files and LD matrices computed using plink with parameter --r square. These sentinel regions were then used to run FINEMAP with the following parameters: --sss --n-causal-snps 10.

#### Fine-mapped Variant Annotation

Fine-mapped variants were annotated using the Ensembl VEP and the Activity-by-Contact genome-wide enhancer maps [90,119]. A credible set of fine-mapped variants was defined as variants with posterior inclusion probability greater than 0.9. This resulted in a fine-mapped set of 529 variants. Credible set variants were annotated using Ensembl VEP for functional sequence type. ABC enhancer maps from CD34+ hematopoietic stem cells were overlapped with the credible set variants. Enrichment of fine-mapped variants within CD34+ enhancers was assessed by comparing the proportion of credible set variants within the ABC CD34+ enhancers to the proportion of variants with a posterior inclusion probability less than 0.1 within the ABC CD34+ enhancers, using a Chi-squared test."

